# Time Course of COVID-19 epidemic in Algeria: Retrospective estimate of the actual burden

**DOI:** 10.1101/2020.06.16.20132746

**Authors:** Mohamed Hamidouche, Nassira Belmessabih

## Abstract

Since December 2019, the five continents have been incrementally invaded by SARS-CoV-2. Africa is the last and least affected to date. However, Algeria is among the first countries affected since February 25, 2020. In order to benefit from its experience in the least affected countries, this study aims to describe the epidemic’s current situation and then retrospectively estimate its real burden.

As a first part of the study, we described the epidemic’s indicators as; the cumulative and daily reported cases and deaths, and we computed the R0 evolution. Secondly, we used the New York City cases-fatality rate standardized by Algerian age structure, to retrospectively estimate the actual burden.

We found that reported cases are in a clear diminution, but, the epidemic epicentre is moving from Blida to other cities. We noted a clear peak in daily cases-fatality from March 30, to April 17, 2020, Fig. 3, due to underestimating the actual infections of the first 25 days. Since May 8, 2020, the daily R0 is around one, Fig. 4. Moreover, we noticed 31% reduction of its mean value from 1,41 to 0,97 between the last two months. The Algerian Age-Standardized Infection Fatality Rate we found is 0,88%. Based on that, we demonstrated that only 1,5% of actual infections were detected and reported before March 30, and 20% after March 31, Fig. 5. Therefore, the actual infections burden is currently five times higher than reported. At the end, we found that at least 0,2 % of the population have been infected until May 27. Consequently, the acquired herd immunity to date is therefore not sufficient to avoid a second wave.

We believe that, the under estimation of the epidemic’s actual burden is probably due to the lack of testing capacities, however, all the indicators show that the situation is currently controlled. This requires more vigilance for the next weeks during the gradual easing of the preventive measures.

## Introduction

All world countries are facing the same threat, the COVID-19 pandemic caused by SARS-CoV-2 infections. Since the first outbreak in Wuhan, China, on December 2019 (Chaolin, et al., 2020; Tan, et al., 2020), the virus spread has reached incrementally the five continents (7,7 million reported cases and more than 428 thousand deaths as of June 14, 2020) (JHU, 2020). First, the pandemic has reached neighbouring Chinese countries in South-East Asia, afterwards, by the end of January 2020, first single cases were reported in the United States (23 January), Brazil (25 February), and in Europe (France: 24 January; Germany: 27 January; Italy and United Kingdom: 31 January; and Spain: 1 February) (Hanns, Michael, & Kathrin, 2020).

Despite that the most of nations have decided to implement a preventive strategy based on a containment, unfortunately, some of them have been very affected by the epidemic, while some others seem to be less affected.

Africa is the last and the least affected continent by the COVID-19 pandemic, the first confirmed case was in Egypt on February 14, 2020. Until June 14, 2020, more than 226 thousands cases and 6070 deaths were reported in the whole continent, and the most affected nations are; South Africa (reported cases: 65736 / reported deaths: 1423), Egypt (42980/1484), Nigeria (15181/399) Ghana (11118/48) and Algeria (10810/760) (AFRO.WHO 1, 2020; Africa CDC, 2020; AFRO.WHO 2, 2020).

In Algeria, the first imported case of COVID-19 was reported on February 25, 2020, when an Italian national tested positive in the southern of the country (Ouargla). On March 1, 2020, the main COVID-19 outbreak began in the northern of Algeria (Blida), when two cases have been confirmed positive after contact with two Algerian nationals residing in France (Algerian Ministry of Health, 2020).

The question that arises, is the data available actual or just the top of the iceberg? Certain hypothesis have been put forwards to explain why the catastrophic forecast scenarios have not been shown so far in Africa, there are three major assumptions; i. the heterogeneity of the population (Gomes, et al., 2020; Lewis, 2020), ii. the cross-protection with other coronaviruses (Angkana, et al., 2020; Lewis, 2020) and iii. the young population in Africa (Fontanet, et al., 2020; Lewis, 2020). However, it is still possible to see an increase in the epidemic in the coming weeks or months (AFRO.WHO 1, 2020). For this reason, it is important to understand the epidemic’s actual burden and evolution in the most affected African countries, thus, their experiences serve as an example to better adapt the implemented preventive strategies in other countries that are at the beginning of the epidemic.

For this purpose, this study aims to describe the current situation of the COVID-19 epidemic in Algeria, then after, based on its indicators, estimate retrospectively its actual burden.

## Methods

In order to describe the current situation of the epidemic in Algeria, we first presented; the reported infections, the cases-fatality and the evolution of daily basic reproduction number (R0). Afterward, we retrospectively estimated the actual burden of COVID-19 epidemic in terms of actual infections, the proportion of reported infections (detection capabilities), and then, estimated the ratio of the population who had already been infected by SARS-CoV-2 in Algeria (the actual cumulative incidence) as of June 14, 2020.

### Epidemiological data

We retrieved information on daily COVID-19 cases and cases-fatality of the ongoing epidemic in Algeria from official reports of governmental institutions and official media in Algeria (Algerian Ministry of Health, 2020; APS 1, 2020).

### The epidemic presentation and indicators estimations

In the descriptive part of the study, we presented graphically the reported daily and cumulative COVID-19 cases and deaths (Fig. 1, Fig. 2, and Fig. 3). In addition, based on reported and confirmed cases with RT-PCR (Algerian Ministry of Health, 2020), we computed the daily R0 and its evolution using the method described in a previous studies (Hamidouche 1, 2020; Hamidouche 2, 2020), while we used the serial interval value SI=4.4 days (Fig. 4) (Chong, et al., 2020).

**Fig. 1.**
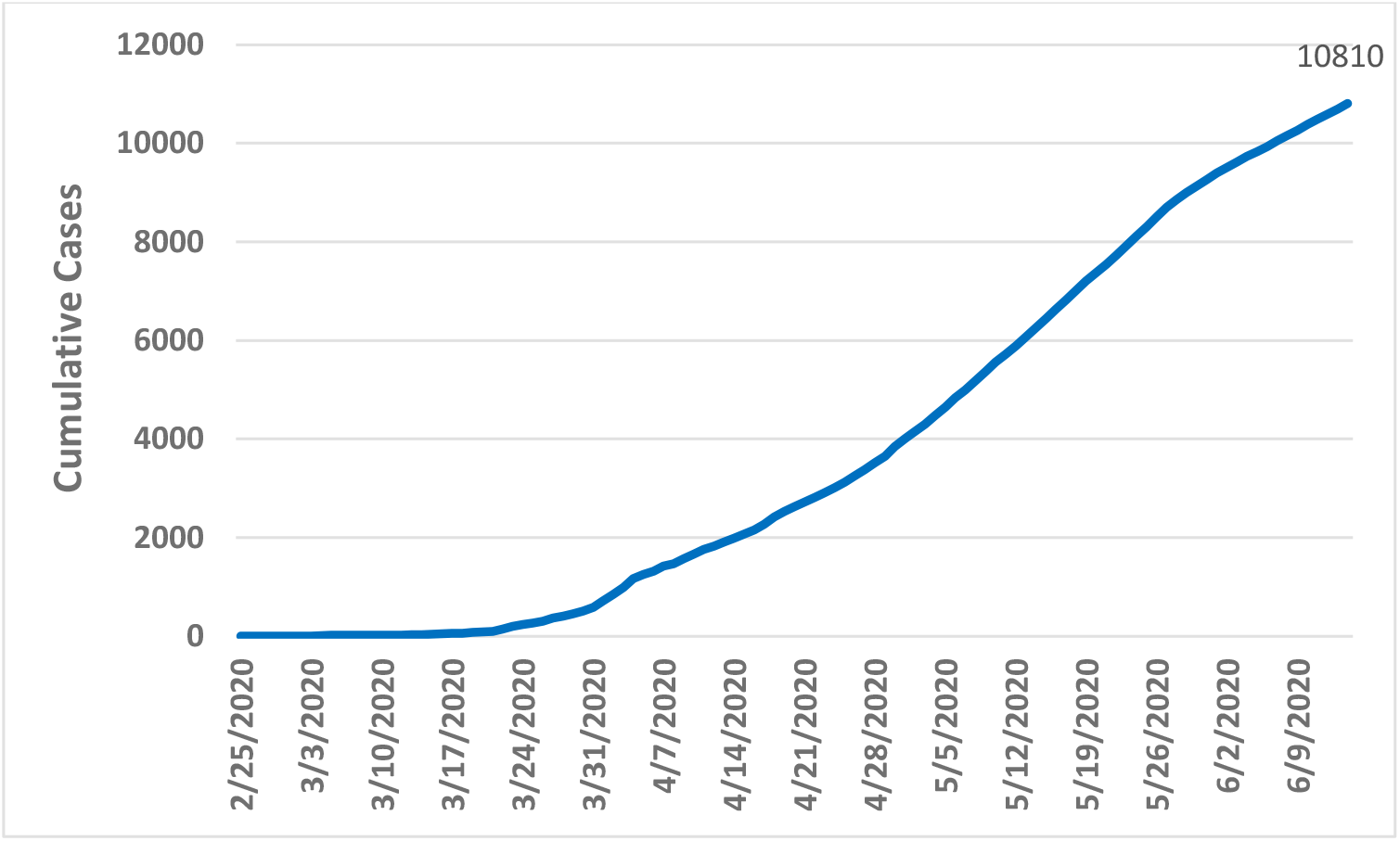
Reported cumulative cases of COVID-19 in Algeria.

**Fig. 2.**
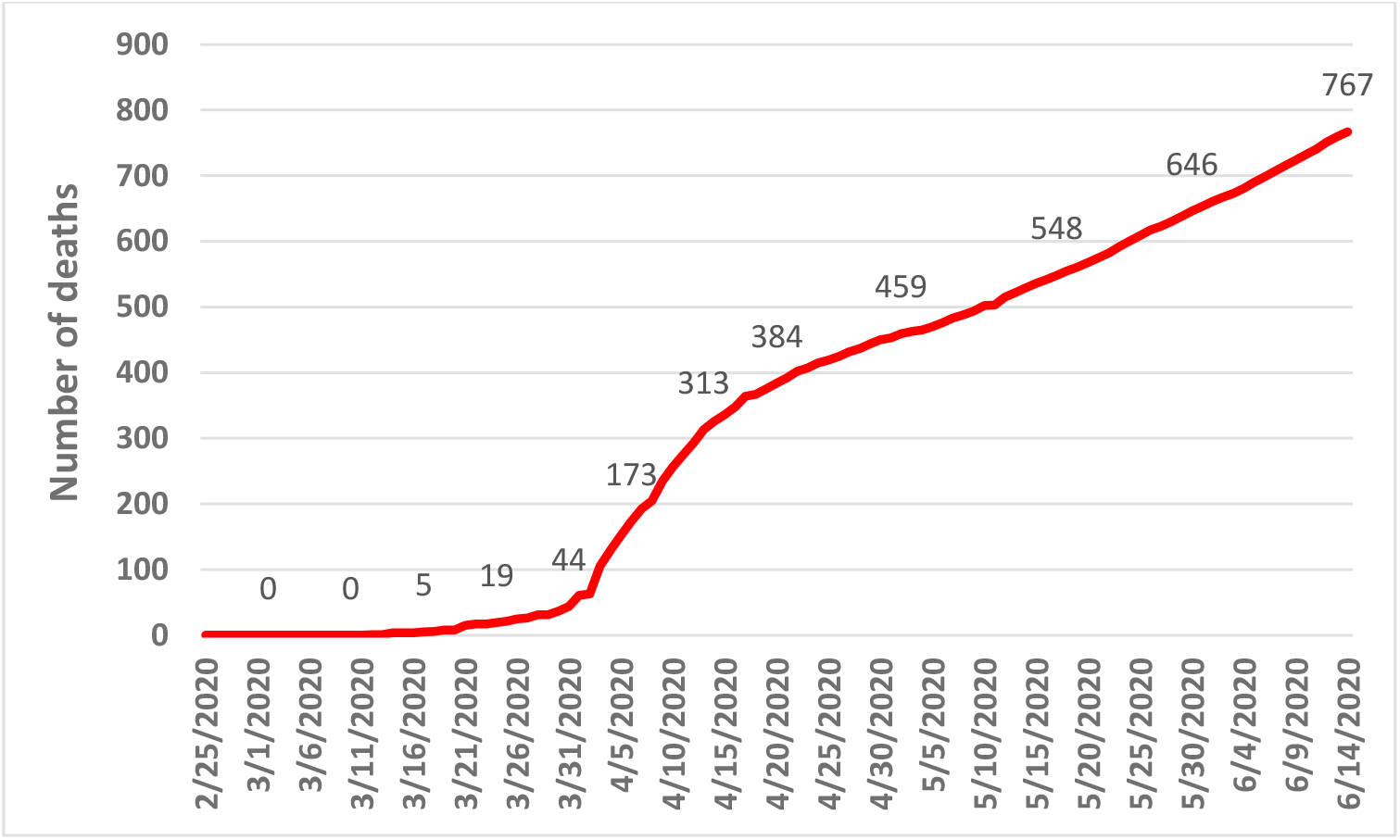
Reported cumulative deaths of COVID-19 in Algeria.

**Fig. 3.**
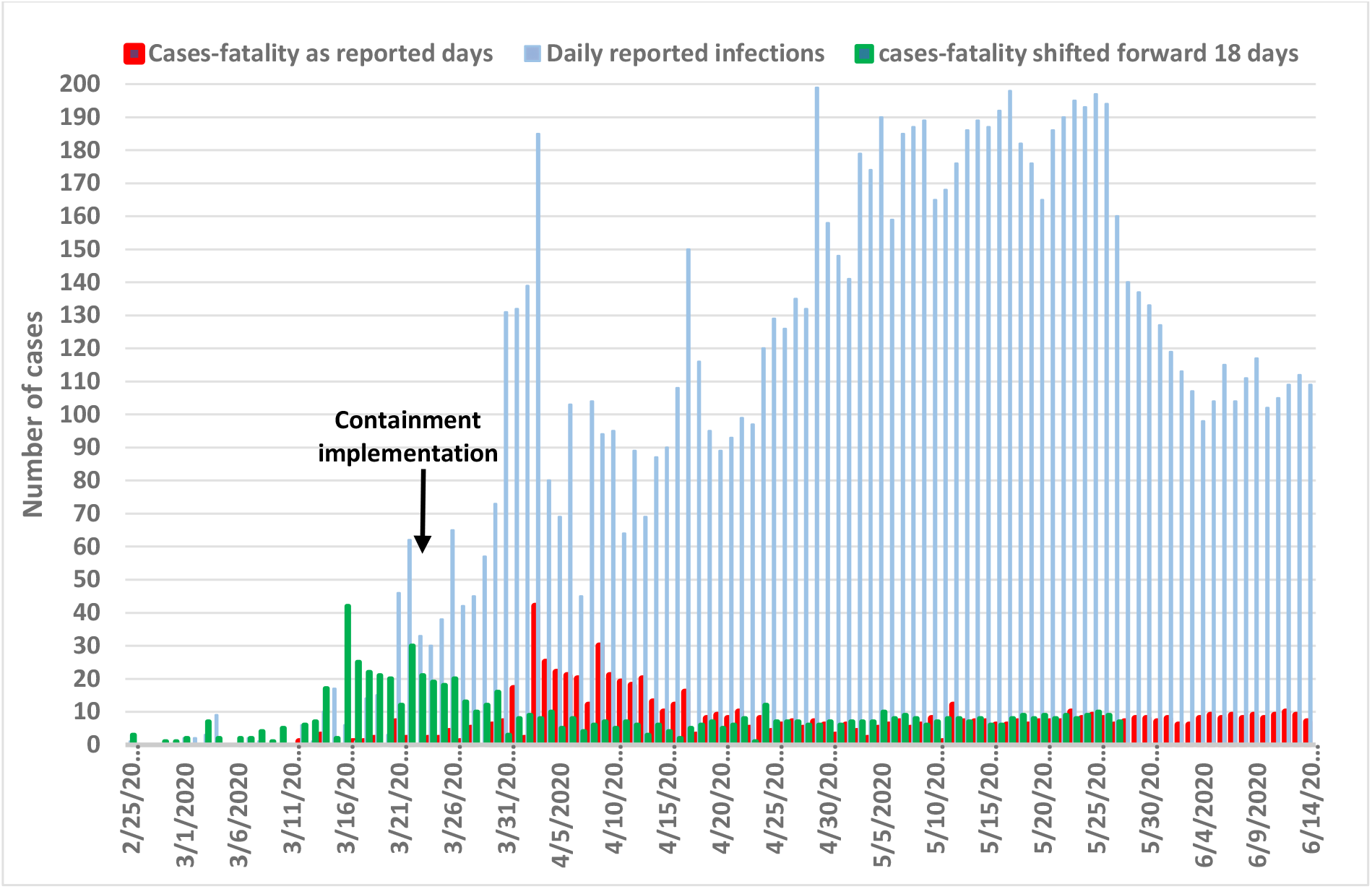
Daily reported COVID-19 cases (blue), and cases-fatality (red) and cases-fatality shifted forwards 18 days as duration from onset of symptoms to death (green).

**Fig. 4.**
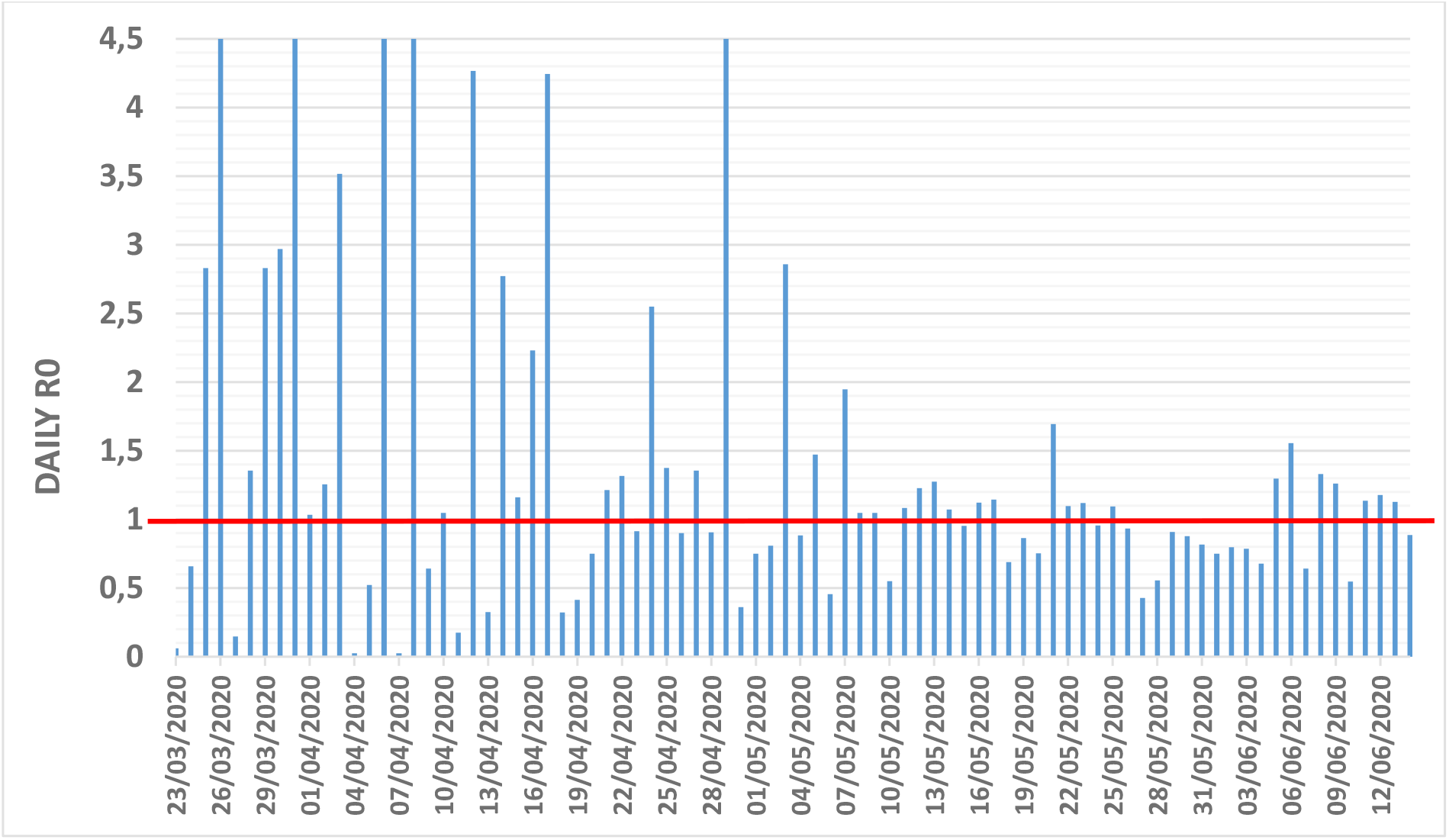
Daily R0 evolution of COVID-19 epidemic in Algeria.

In order to reveal if the numbers of infections were underestimated or not, we overlapped the daily reported infections and cases-fatality, then, we shifted the latter forwards 18 days on the graph (Fig. 3) to take into account the mean duration from onset of symptoms to death (17,8 days) (95% 16,9-19,2) (Robert, et al., 2020). Due to the low testing rate in Algeria (6500 tests) (154 test/M population) until April 13, 2020, among them 1907 test (29%) were positive (Hamidouche 2, 2020; El Watan, 2020), we consider that the majority of COVID-19 cases are reported after the symptoms onset. For this reason, we did not add the median incubation period of 5.1 days (95% CI 4,5-5,8 days) (Lauer, Grantz, & Bi, 2020) to the 18 days of symptoms onset.

The second part of the study was devoted to estimating the actual burden of the COVID-19 epidemic in Algeria. It is known that the case-fatality rate vary from a country to another, from 1 % to 16% (worldometers, 2020). This variation is conceivable to be related to several factors, for instance, testing capacities, vulnerable group’s rates among the population and medical care’s capacity and quality in the concerned country (Dong-Hyun, Young, & Jin-Young, 2020).

To estimate the most accurate Infection Fatality Rate (IFR) to date for COVID-19 epidemic in Algeria, we decided to use the New York City IFR (NYC-IFR) to compute the Algerian Age-Standardized Infection Fatality Rate (AS-IFR) (Dong-Hyun, Young, & Jin-Young, 2020). We chose to use the NYC-IFR because of its rigorous calculation method practiced on real data and not on predictions. The numerator of the NYC-IFR was based on data of deaths provided by NYC (18282 deaths) (NYC, 2020), plus, the excess deaths analysis by the CDC (5293 more deaths above expected seasonal baseline levels) (CDC 1, 2020). And the denominator of NYC-IFR that represents the number of the already infected population is the result of the New York State antibody study (1,7 million people (19.9%) in NYC had been infected with SARS-CoV-2 as of May 1, 2020) (New York State, 2020). The result was NYC-IFR=1.4% (worldometers, 2020).

We used this value (NYC-IFR=1.4%), to calculate the AS-IFR, which in turn was used to retrospectively estimate the actual infections during the COVID-19 epidemic in Algeria (the fatality-based retrospectively estimated infections), since its beginning on March 25, until June 14, 2020.

### Statistical analysis and software

This study was carried out using Excel 2013 and STATA/IC 15 software. We also used the Alg-COVID-19 Model established in a previous study to estimate the R0 (Hamidouche 2, 2020; Hamidouche 1, 2020).

## Results

The number of new confirmed cases is raising since the first case declaration on February 25, 2020, consequently, the deaths too (Fig. 1, Fig. 2). As it was warned in advance (Hamidouche 2, 2020), we see that the main outbreak is moving from Blida (1372 cumulative cases) to other cities like; Algiers (1212 cumulative cases), Setif (756 cumulative cases) and Oran (678 cumulative cases) (data of June 14, 2020) (Algerian Ministry of Health, 2020).

Distinctly, there was an exponential phase in daily reported infections until April 29, 2020, then they stabilized between 150 and 198 cases/day for 25 days (from April 29 to May 25, 2020), then after, we noticed a clear diminution. Although, we can see some oscillation and unexpected peaks over the curve (Fig. 3).

Regarding the daily cases-fatality, we noted a clear peak from March 30, to April 17, 2020, (Fig. 3). After shifting forwards, the bar chart of the daily cases-fatality over the reported daily COVID-19 cases 18 days to match it approximately with the dates of infections that cause them. We noticed that, the actual infections have been underestimated over the period of the noted peak, the overlapped reported cases-fatality of at least the first 25 days of the epidemic were very high in comparison with the reported infections (Fig. 3).

Based on the reported cases, the computed daily R0 evolution since March 23, 2020, was unstable and presented sometimes values higher than >4,5, however, since, May 8, 2020, the daily R0 value was the mostly around 1 (Fig. 4). Moreover, we noticed 31 % reduction of the average R0 of the previous month (April 16, to May 15, 2020) from 1,41 (95% CI 1,26-1,56) to 0.97 (95% CI 0,93-1,0) in the last month (May 16, to June 14, 2020).

As reported in the methods section, based on NYC-IFR=1,4%, the Algerian Age-Standardized Infection Fatality Rate (AS-IFR) that we found is 0,88%. We used this value to compute the actual daily infections retrospectively. Thus, after taking into account 18 days to consider the mean duration from onset of symptoms to death, we presented graphically (Fig. 5) the fitting between daily reported infections and fatality-based retrospectively estimated infections. We have seen that the gap between the estimated infections and the reported infections was considerable between March 12 and March 30, 2020, knowing that the epidemic has started in the main outbreak in Blida, on March 01, 2020. We found that, only an average of 1,5% of infections was detected and reported among the total estimated infections (from February 25 to March 30, 2020), and an average of 20% for the duration between March 31 and June 14, 2020 (Fig. 5). So, based on AS-IFR=0,88 we can deduce that the real burden of infection is therefore five times higher than detected and reported.

**Fig. 5.**
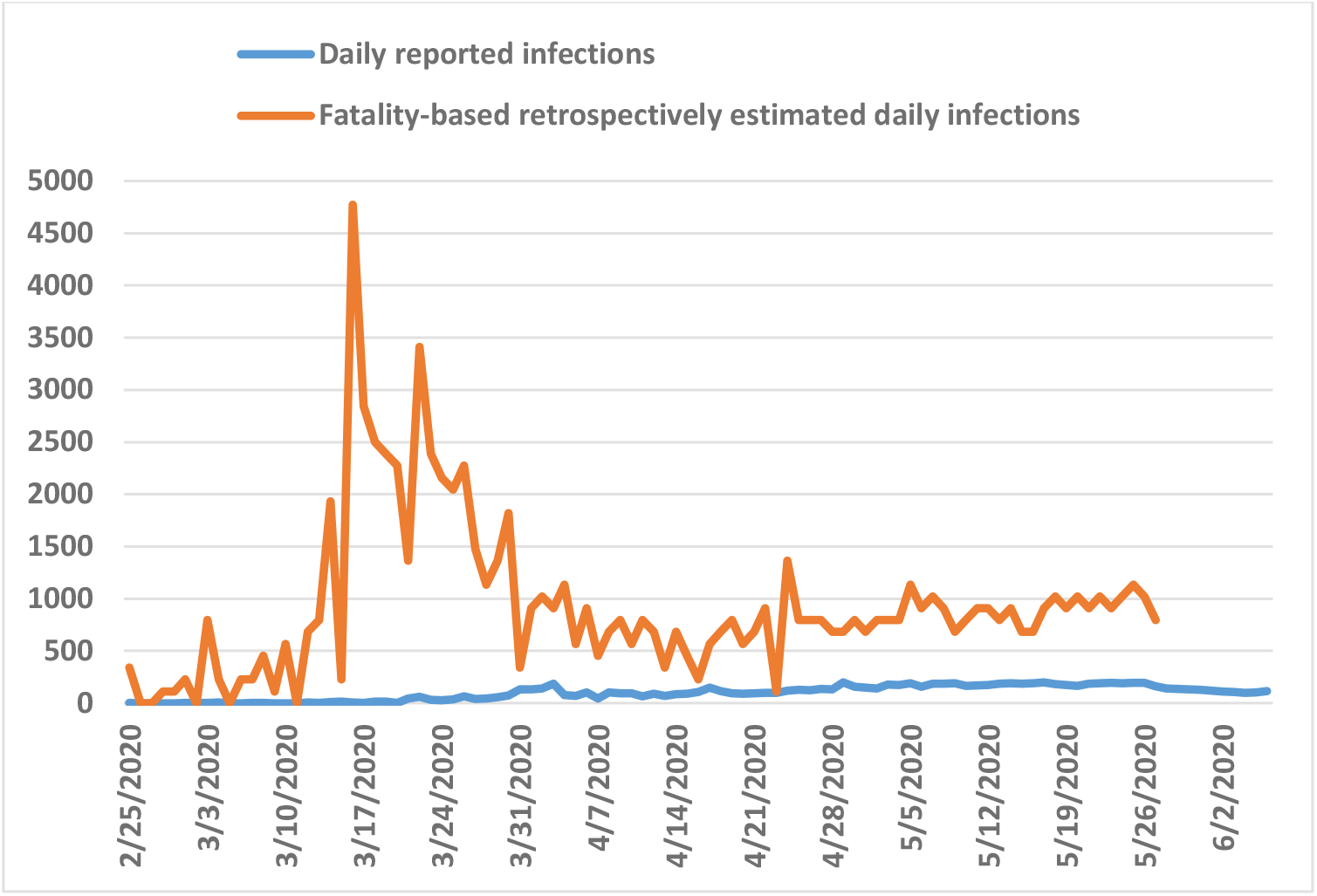
Fitting between COVID-19 daily reported cases (blue), and fatality-based retrospectively estimated infections (AS-IFR=0,88%)(red), taking into account 18 days as duration from onset of symptoms to death.

At the end, the comparison between cumulative reported (detected) cases and estimated (actual) infections based on AS-IFR=0,88, has revealed that, at least 87045 persons have been infected until May 27, 2020, which represent 0,2 % of the Algerian population (Fig. 6).

**Fig. 6.**
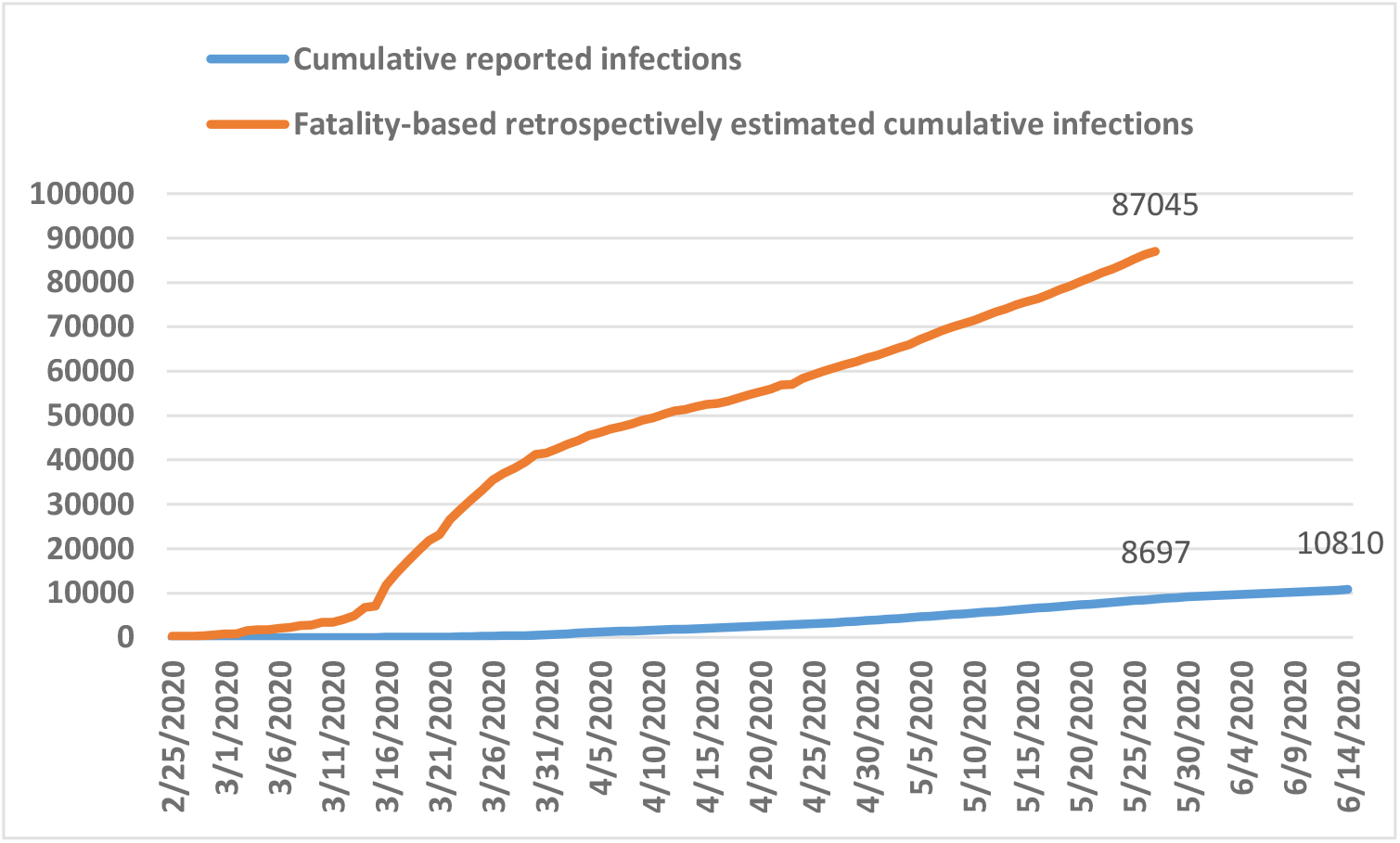
COVID-19 reported cases (blue) and fatality-based retrospectively estimated cumulative cases (AS-IFR=0,88%) (red), taking into account 18 days as duration from onset of symptoms to death.

## Discussion

The unexpected variations, and increases in reported infections, such as the peak reported from March 31 to April 03, 2020 (Fig. 3), could be due to an artifact resulting from the notification of cases by the newly recruited RT-PCR testing laboratories. As more than 20 laboratories have been deployed across the country since the start of the epidemic, which has doubled the testing capacity from 200 to 400 test/day (Algerian Ministry of Health, 2020; APS 2, 2020).

The clear peak in reported cases-fatality between March 30, and April 17, 2020, is probably due to a considerable not detected increase in daily infections about 18 days earlier (the duration of the onset of symptoms until death) (Robert, et al., 2020), or rather, since the beginning of the epidemic on February 25, 2020 (Fig. 3). After shifting forwards the case fatality bar chart 18 days over the reported daily COVID-19 cases (to match the dates of infections that cause them). The underestimation of actual infections that is presented clearly on Fig. 3, is obviously due to the low capacity of testing at the beginning of the outbreak (El Watan, 2020). Furthermore, the assumption of an earlier introduction (before February 25, 2020) of the virus into the country is possible, but, without evidence so far, as the cases tracing results are not publicly available.

As a retrospective analysis, we can say that, the noted peak of cases-fatality (from March 30, 2020 to April 17, 2020) could have been avoided if the preventive measures had been implemented by March 11, 2020 at the latest (Fig. 3). Especially, the total containment implementation in Blida as a main outbreak, knowing that, at that time, the majority of casualties were located there (Algerian Ministry of Health, 2020).

Regarding the R0 evolution since March 23, 2020, it was very fluctuating, and sometimes its value was higher than 4,5. Nevertheless, the value has stabilized around 1 over time (Fig. 4). This is very likely to be related to the reporting variation at the beginning, then, to the effectiveness of the preventive measures implemented since March 23, 2020 (Algerian Ministry of Health, 2020; Hamidouche 2, 2020). From May 8, 2020, the daily R0 was mostly around 1, its reduction of 31 % between the last two months (from 1,41 (95% CI 1,26-1,56) (April 16 to May 15) to 0.97 (95% CI 0,93-1,0) (May 16 to June 14)) is a good sign of good control actions (Fig. 4).

However, the confidence interval of the last value of R0 includes the value 1, this is not enough to reduce the incidence so as to put an end to the epidemic. To achieve this objective, it must be kept under the value of 1, which is difficult to reach especially if the undetected infections rate remains high (according to our results above, the undetected infections represent around 80% of actual infections in Algeria). For comparison, in Wuhan, china, the estimated R0 of COVID-19 after two months of the lockdown ranged from 1,66 (95% CI 0,72-2,87) (Ku, Ng, & Lin, 2020).

On the other hand, this indicator (R0) reflects that the COVID-19 epidemic is under control. This inference is confirmed by the stability, then, the recent decline of the daily reported infections curve (Fig. 3), and more importantly, the low number of COVID-19 patients that are sojourning in intensive care units across the country. Only 32 cases on daily average are under intensive care during the last 30 days (from May 16, to June 14, 2020). According to the health minister declaration, the use of intensive care capacity did not exceed 17% since the beginning of the epidemic (Algerian Ministry of Health, 2020).

Worldwide, it is difficult to detect all infections during an epidemic, and some epidemic indicators are more reliable than others, because they result from the infections and they are inevitable, for instance, the cases-fatality, the intensive care access, and the hospitalizations, so that, these are the most used parameters to oversight an epidemic. For these reasons, and for data availability, we only used the infection fatality rate to estimate the actual burden of the COVID-19 epidemic in Algerian.

We standardized the IFR by age, because of the difference in age structure of affected population between countries may guide to the true burden of COVID-19 in terms case fatality rate, and it is useful in comparing between the countries (Dong-Hyun, Young, & Jin-Young, 2020). It is agreed that age is the most associated factor to COVID-19 deaths, and most of the underlying diseases are positively correlated with age (CDC 2, 2020; Chen, et al., 2020; Guan, et al., 2020). So, we believe that, the found result of Algerian Age-Standardized Infection Fatality Rate (AS-IFR=0,88%) is more accurate than the reported case-fatality rate (7% until June 14, 2020), because, the latter is based only on reported cases as a denominator. In addition, the found value is in accordance with the IFR that are estimated worldwide between 0,5% and 1,5%, for instance; the IFR in France 0.7% (Henrik, et al., 2020), in Italy 1.31% (Gianluca & Paradisi, 2020), and in China 0,66% (Robert, et al., 2020).

By considering 18 days of the mean duration from symptoms onset to death, and based on AS-IFR=0,88%, we believe that the fatality-based retrospectively estimated infections are the more accurate possible.

As reported in the result section, the infections detection capacities have increased from 1.5% to 20% recently, probably due to the increase in testing capacities from 200 to 400 tests per day (APS 2, 2020), and to the experience gained by the epidemic monitoring system (Fig 5).

The cumulative estimated infections demonstrated that the actual infections are five time higher than detected and reported, thus, at least 87045 persons have been infected after three months and 20 days (by June 14, 2020), which represents around 0,2% of the Algerian population (Fig. 6), however, this can vary from a region to another.

This finding (0,2%) seems very low compared to the results of other studies, for instance, a modelling study has shown that 4.4% of the French population had been infected by May 11, 2020, (four and a half months after the first case reported on December 27, 2019) (Henrik et al., 2020). Also, results from a serological study show that 19.9% of the New York population had anti-SARS-CoV-2 antibodies after three months and 15 days from the date of the first case (March 1, 2020) (NYC, 2020). The previously mentioned examples show that, the cumulative incidence of infections in Algeria, may be higher than 0,2%, even if it is the case, the herd immunity appears insufficient to avoid a second wave if all control measures are released by the end of the lockdown. It is also possible that the cumulative incidence is low because of the strong measures that the government has implemented since March 23, 2020 (Hamidouche 2, 2020).

Our results may have some limitations, the R0 estimation is based on the reported cases, also, the found AS-IFR may not be very precise due to the age-standardization method we used or other factors related to countries specifications, consequently, the actual burden of infections might vary around the found estimations. Nevertheless, we believe that our results are very useful in clarifying the vision for decision-makers, and can help to understand the current situation of the epidemic in Algeria, thus, it can be used to adjust the after containment plan that is already in place since June 07, 2020. However, only a good serological study, based on a representative sampling and a good reliable serological tests can reveal the actual burden of the epidemic in Algeria.

## Conclusion

Undoubtedly, the preventive strategy that have been implemented on March 23, 2020, was effective and resulted in avoiding thousands of infections and hundreds of deaths, and its effects are reflected on the results of this and a previous one (Hamidouche 2, 2020).

However, underestimating the SARS-CoV-2 infections that we noticed between March 12 and March 30, 2020 (Fig. 5), and, as consequences, the peak of cases-fatality observed 18 days later (from March 30 to April 17, 2020) had caused additional causalities (Fig. 3).

Despite Algeria was among the first and most affected countries by the epidemic in Africa, the SARS-CoV-2 seroprevalence remains very low. This should be considered for any easing of control actions to avoid new outbreaks. Therefore, it is necessary to implement and enforce a mitigation measure accompanying the gradual release of containment, especially, actions that have proven to be effective. For example, good hygiene habits, physical distancing and wearing medical mask that has been reported to reduce the risk of viral transmission by 85% (Renyi Zhang, Annie, Yuan, & Mario, 2020; Derek, et al., 2020). The adoption of the previously mentioned actions by the stakeholders and ordinary people remain the best solution to leave the lockdown and alleviate the restrictions without major risk.

Finally, this study shows that, the COVID-19 epidemic in Algeria is currently under control, and the evolution of all its parameters, such as; the daily reported infections, the basic reproduction number, the number of deaths per day and the daily cases in the intensive care units, lead to the same conclusion. Bearing in mind that the hypothesis (cross-protection (Angkana, et al., 2020), infected children as a dead-end (Fontanet, et al., 2020) and the population heterogeneity (Gomes, et al., 2020)) may have a positive influence on the controlled situation.

## Data Availability

All used data is well mentioned with DOI and links in the references part

## Notes

### Competing Interest Statement

The authors have declared no competing interest.

### Funding Statement

No funding was allocated to this study

